# Mental Health Conditions in Lesbian, Gay, Bisexual, Transgender, Queer and Asexual Youth in Brazil: a call for action

**DOI:** 10.1101/2021.06.24.21259474

**Authors:** Tauana Terra, Julia L. Schafer, Pedro M. Pan, Angelo Brandelli Costa, Arthur Caye, Ary Gadelha, Eurípedes Miguel, Rodrigo A. Bressan, Luis A. Rohde, Giovanni A. Salum

## Abstract

Lesbian, Gay, Bisexual, Transgender, Queer and Asexual (LGBTQA+) youth have a greater odds of experiencing stressful life events like discrimination and violence when compared to their cisgender heterosexual peers, which can lead to mental health problems. We aimed to estimate the prevalence of mental disorders among LGBTQA+ youths from the 3rd wave of the Brazilian High-Risk Cohort for Psychiatric Disorders (n= 1,475). Mental disorders were assessed using the Brazilian version of the Development and Well-Being Behavior Assessment. Sexual orientation, gender identity and biological sex were assessed using specific questions of a self-report confidential questionnaire. Data were analyzed using sampling weights to account for attrition and our oversampling design. The mean age was 18.14 years (SD= 2.00) and 15.18% of the sample described themselves as LGBTQA+ (n=221). The LGBTQA+ group presented higher rates of anxiety disorders (30.14% vs. 13.37%; OR= 3.66; 95%CI: 2.82 - 4.75; p<0.001), depressive disorders (27.75% vs. 15.34%; OR= 2.51; 95%CI: 1.92 - 3.27, p<0.001) and post-traumatic stress disorder (4.98% vs. 2.25%, OR= 4.21, 95%CI: 2.54 - 6.96, p<0.001), if compared with the cisgender heterosexual group. No difference was found for conduct disorders (2.97% vs. 5.21% OR= 0.81; 95%CI: 0.39 - 1.69; p=0.577) or attention deficit hyperactivity disorder (5.92% vs. 3.28%, OR= 1.29; 95%CI: 0.74 - 2.25; p=0.361). Our results elucidate the mental health disparities between LGBTQA+ people and cisgender heterosexuals in Brazil. It highlights the need to promote the inclusion of this population in policy formulation and to support actions to mitigate and prevent the suffering and mental health problems related to sexual orientation and gender identity.

## Introduction

Lesbian, Gay, Bisexual, Transgender, Queer and Asexual (LGBTQA+) youth have a greater odds of experiencing stressful life events like discrimination and violence when compared to their cisgender heterosexual peers,^1^ which can lead to mental health problems. Nevertheless, data from community surveys conducted in low-and middle-income countries, where levels of violence against sexual minorities tend to be higher,^2^ is scarce.^3,4^ Here we estimate the prevalence of mental disorders among LGBTQA+ youths from the 3rd wave of the Brazilian High-Risk Cohort for Psychiatric Disorders (BHRCS).^5^ Briefly, 2,511 participants were assessed in baseline (38% randomly selected; 62% selected as high risk according to screening child and family psychiatric symptoms; for details see^3^). From these a total of 1,905 was re-evaluated at wave 3 (75.86% retention) and from those 1,475 answered all questions about sexual orientation and gender identity in the confidential protocol.

## Methods

Mental disorders were assessed using the Brazilian version of the Development and Well-Being Behavior Assessment (DAWBA). This structured interview was administered to parents by trained lay interviewers and by psychologists to study participants. Responses were then evaluated by certified psychiatrists, which confirmed, refuted, or altered the initial computerized diagnosis. Diagnoses used for data analysis were any anxiety disorder (separation anxiety, panic, social anxiety, or generalized anxiety disorder), any depressive disorder (major depression, or depression not otherwise specified), any conduct disorder (oppositional defiant disorder or conduct disorder), post-traumatic stress disorder (PTSD) and any attention deficit hyperactivity disorder (ADHD) and any disorder (any of the aforementioned disorders).

Sexual orientation, gender identity and biological sex were assessed using explanations followed by specific questions in a self-report confidential questionnaire. Sexual orientation: “Sexual orientation refers to the emotional, physical and sexual attraction we feel for other people. What is your sexual orientation?”, which have the following response options: 1) Heterosexual; 2) Bisexual; 3) Homosexual; 4) Asexual; 5) Other. Gender identity: “Gender Identity concerns the way you feel, perceive yourself and the way you would like other people to recognize you. That is, it is the gender that you identify yourself with. How do you identify yourself?”, which have the following response options: 1) Woman; 2) Man; 3) Trans woman; 4) Trans man; 5) Travesti; 6) Other; 7) I don’t know. Biological sex: “Gender determined at birth refers to how identified when we were born. What is your gender identified at birth?”, which have the following response options: 1) Woman; 2) Man. Data were analyzed using sampling weights to account for attrition and our oversampling design. Weights were constructed to represent the sample randomly selected at the BHRC baseline, using the Covariate Balancing Propensity Score package^6^ with symptoms collected at baseline as input for the propensity score weights. The association between sexual orientation/gender identity and mental health conditions were identified using binary logistic regression. For logistic regressions extreme weights were trimmed below the 5th and above the 95th percentile.^7^

## Results

The mean age of participants was 18.14 years (SD= 2.00) and 51.74% were identified at birth as male. According to the Brazilian Socioeconomic Criteria most people (62.23%) belonged to category C, 23.59% to A/B (the wealthiest), and 14.18% to D/E category (the poorest). A total of 15.18% of the sample described themselves as LGBTQA+ (n=221). Sexual orientation: Lesbian= 12.22%, Gay= 12.67%, Bisexual = 63.35%, Asexual = 8.14% and Pansexual = 1.81%. Gender Identity: Transgender = 4.07 %, Agender = 0.45% and Non-binary = 0.90%. Additionally, 0.45% identified as Queer regardless of sexual orientation and gender identity. The prevalence of at least one mental disorder was 51.95% in LGBTQA+vs. 32.70% in cisgenders heterosexuals (OR= 2.25; 95%CI: 1.78 - 2.84; p<0.001). The LGBTQA+ group presented higher rates of anxiety disorders (30.14% vs. 13.37%; OR= 3.66; 95%CI: 2.82 - 4.75; p<0.001), depressive disorders (27.75% vs. 15.34%; OR= 2.51; 95%CI: 1.92 - 3.27, p<0.001) and PTSD (4.98% vs. 2.25%, OR= 4.21, 95%CI: 2.54 - 6.96, p<0.001), if compared with the cisgender heterosexual group. No difference was found for conduct disorders (2.97% vs. 5.21% OR= 0.81; 95%CI: 0.39 - 1.69; p=0.577) or ADHD (5.92% vs. 3.28%, OR= 1.29; 95%CI: 0.74 - 2.25; p=0.361).

## Discussion

Our results elucidate the mental health disparities between LGBTQA+ people and cisgender heterosexuals in Brazil. Minority stress theory^1^ has provided a foundational framework for understanding LGBTQA+ mental health disparities such as chronic stressors related to their stigmatized identities, including victimization, prejudice, and discrimination which need further investigation in this sample. These distinct experiences, in addition to everyday or universal stressors, disproportionately compromise the mental health and well-being of LGBTQA+ adolescents, and seems to persist until adulthood.^7^ This study provides a significant contribution to the understanding of mental health disparities between LGBTQA+ youth and cisgenders heterosexuals in developing countries. It highlights the need to promote the inclusion of this population in policy formulation and to promote programs, plans, and actions to mitigate and prevent the suffering and mental health problems related to sexual orientation and gender identity.

## Data Availability

Data is available under request.

## Acknowledgements

This work is supported by the National Institute of Developmental Psychiatry for Children and Adolescents, a science and technology institute funded by Conselho Nacional de Desenvolvimento Científico e Tecnológico (CNPq; National Council for Scientific and Technological Development; grant numbers 573974/2008-0 and 465550/2014-2) and Fundação de Amparo à Pesquisa do Estado de São Paulo (FAPESP; Research Support Foundation of the State of São Paulo; grant number 2008/57896-8 and 2014/50917-0).

